# Effectiveness of the Coronavirus Disease 2019 (COVID-19) Bivalent Vaccine

**DOI:** 10.1101/2022.12.17.22283625

**Authors:** Nabin K. Shrestha, Patrick C. Burke, Amy S. Nowacki, James F. Simon, Amanda Hagen, Steven M. Gordon

## Abstract

**Background:** The purpose of this study was to evaluate whether a bivalent COVID-19 vaccine protects against COVID-19.

**Methods:** Employees of Cleveland Clinic in employment when the bivalent COVID-19 vaccine first became available, were included. Cumulative incidence of COVID-19 over the following 26 weeks was examined. Protection provided by vaccination (analyzed as a time-dependent covariate) was evaluated using Cox proportional hazards regression, with change in dominant circulating lineages over time accounted for by time-dependent coefficients. The analysis was adjusted for the pandemic phase when the last prior COVID-19 episode occurred, and the number of prior vaccine doses.

**Results:** Among 51017 employees, COVID-19 occurred in 4424 (8.7%) during the study. In multivariable analysis, the bivalent vaccinated state was associated with lower risk of COVID-19 during the BA.4/5 dominant (HR, .71; 95% C.I., .63-.79) and the BQ dominant (HR, .80; 95% C.I., .69-.94) phases, but decreased risk was not found during the XBB dominant phase (HR, .96; 95% C.I., .82-.1.12). Estimated vaccine effectiveness (VE) was 29% (95% C.I., 21%-37%), 20% (95% C.I., 6%-31%), and 4% (95% C.I., -12%-18%), during the BA.4/5, BQ, and XBB dominant phases, respectively. Risk of COVID-19 also increased with time since most recent prior COVID-19 episode and with the number of vaccine doses previously received.

**Conclusions:** The bivalent COVID-19 vaccine given to working-aged adults afforded modest protection overall against COVID-19 while the BA.4/5 lineages were the dominant circulating strains, afforded less protection when the BQ lineages were dominant, and effectiveness was not demonstrated when the XBB lineages were dominant.

**Summary:** Among 51017 working-aged Cleveland Clinic employees, the bivalent COVID-19 vaccine was 29% effective in preventing infection while the BA.4/5 lineages were dominant, and 20% effective while the BQ lineages were. Effectiveness was not demonstrated when the XBB lineages were dominant.

## INTRODUCTION

When the original messenger RNA (mRNA) Coronavirus Disease 2019 (COVID-19) vaccines first became available in 2020, there was ample evidence of efficacy from randomized clinical trials [1,2].Vaccine effectiveness was subsequently confirmed by clinical effectiveness data in the real world outside of clinical trials [3,4], including an effectiveness estimate of 97% among employees within our own healthcare system [5]. This was when the human population had just encountered the novel Severe Acute Respiratory Syndrome Coronavirus 2 (SARS-CoV-2) virus, and the pathogen had exacted a high burden of morbidity and mortality across the world. The vaccines were amazingly effective in preventing COVID-19, saved a large number of lives, and changed the impact of the pandemic.

Continued acquisition of mutations in the virus, from natural evolution in response to interaction with the immune response among the human population, led to the emergence and spread of SARS-CoV-2 variants. Despite this, for almost two years since the onset of the pandemic, those previously infected or vaccinated continued to have substantial protection against reinfection by virtue of natural or vaccine-induced immunity [6]. The arrival of the Omicron variant in December 2021, brought a significant change to the immune protection landscape. Previously infected or vaccinated individuals were no longer protected from COVID-19 [6]. Vaccine boosting provided some protection against the Omicron variant [7,8], but the degree of protection was not near that of the original vaccine against the pre-Omicron variants of SARS-CoV-2 [8]. After the emergence of the Omicron variant, prior infection with an earlier lineage of the Omicron variant protected against subsequent infection with a subsequent lineage [9], but such protection appeared to wear off within a few months [10]. During the Omicron phase of the pandemic, protection from vaccine-induced immunity decreased within a few months after vaccine boosting [8].

Recognition that the original COVID-19 vaccines provided much less protection after the emergence of the Omicron variant spurred efforts to produce newer vaccines that were more effective. These efforts culminated in the approval by the US Food and Drug Administration, on 31 August 2022, of bivalent COVID-19 mRNA vaccines, which encoded antigens represented in the original vaccine as well as antigens representing the BA.4/5 lineages of the Omicron variant. Given the demonstrated safety of the earlier mRNA vaccines and the perceived urgency of need of a more effective preventive tool, these vaccines were approved without demonstration of effectiveness in clinical studies.

The purpose of this study was to evaluate whether the bivalent COVID-19 vaccine protects against COVID-19.

## METHODS

### Study design

This was a retrospective cohort study conducted at the Cleveland Clinic Health System (CCHS) in the United States. The study was approved by the Cleveland Clinic Institutional Review Board as exempt research (IRB no. 22-917). A waiver of informed consent and waiver of HIPAA authorization were approved to allow the research team access to the required data.

### Setting

Since the arrival of the COVID-19 pandemic at Cleveland Clinic in March 2020, employee access to testing has been a priority. Voluntary vaccination for COVID-19 began on 16 December 2020, and the monovalent mRNA vaccine as a booster became available to employees on 5 October 2021. The bivalent COVID-19 mRNA vaccine began to be offered to employees on 12 September 2022. This date was considered the study start date.

The mix of circulating variants of SARS-CoV-2 changed over the course of the study. The majority of infections in Ohio were initially caused by the BA.4 or BA.5 lineages of the Omicron variant. By mid-December 2022 the BQ lineages, and by mid-January 2023 the XBB lineages of the Omicron variant, were the dominant circulating strains [11].

### Participants

CCHS employees in employment at any Cleveland Clinic location in Ohio on 12 September 2022, the day the bivalent vaccine first became available to employees, were included in the study. Those for whom age and gender were not available were excluded.

### Variables

Covariates collected were age, sex, job location, and job type categorized into clinical or non-clinical, as described in our earlier studies [5–7]. Institutional data governance rules related to employee data limited our ability to supplement our dataset with additional clinical variables. Subjects were considered pre-pandemic hires if hired before 16 March 2020, the day COVID-19 testing became available in our institution, and pandemic hires if hired on or after that date.

Prior COVID-19 was defined as a positive nucleic acid amplification test (NAAT) for SARS-CoV-2 any time before the study start date. The date of infection for a prior episode of COVID-19 was the date of the first positive test for that episode of illness. A positive test more than 90 days following the date of a previous infection was considered a new episode of infection. Since the health system never had a requirement for systematic asymptomatic employee test screening, most positive tests would have been tests done to evaluate suspicious symptoms. Some would have been tests done to evaluate known exposures or tests for pre-operative or pre-procedural screening.

The pandemic phase (pre-Omicron or Omicron) during which a subject had his or her last prior episode of COVID-19 was also collected as a variable, based on which variant/lineages accounted for more than 50% of infections in Ohio at the time [11].

### Outcome

The study outcome was time to COVID-19, the latter defined as a positive NAAT for SARS-CoV-2 any time after the study start date. Outcomes were followed until 14 March 2023, allowing for evaluation of outcomes up to 26 weeks from the study start date.

### Statistical analysis

A Simon-Makuch hazard plot [12] was created to compare the cumulative incidence of COVID-19 in the bivalent vaccinated and non-vaccinated states, by treating bivalent vaccination as a time-dependent covariate. Individuals were considered bivalent vaccinated 7 days after receipt of a single dose of the bivalent COVID-19 vaccine. Subjects whose employment was terminated during the study period before they had COVID-19 were censored on the date of termination. Curves for the non-vaccinated state were based on data while the bivalent vaccination status of subjects remained “non-vaccinated”. Curves for the bivalent vaccinated state were based on data from the date the bivalent vaccination status changed to “vaccinated”.

Multivariable Cox proportional hazards regression models were fitted to examine the association of various variables with time to COVID-19. Bivalent vaccination was included as a time-dependent covariate [13]. The study period was divided into BA.4/5 dominant, BQ dominant, and XBB dominant phases, depending on which group of lineages accounted for more than 50% of all COVID-19 infections at the time (based on variant proportion data from the Centers for Disease Control and Prevention) [11], and which group of lineages was most abundant in our internal sequencing data. Time-dependent coefficients were used to separate out the effects of the bivalent vaccine during the different phases. The primary model included all study subjects. The secondary model included only those with prior exposure to SARS-CoV-2 by infection or vaccination, and evaluated the effect of bivalent vaccination with inclusion of time since most recent exposure to SARS-CoV-2 by infection or vaccination, to adjust for the effect of waning immunity on susceptibility to COVID-19. The possibility of multicollinearity in the models was evaluated using variance inflation factors. The proportional hazards assumption was checked using log(-log(survival)) vs. time plots. Vaccine effectiveness was calculated from the hazard ratios for bivalent vaccination in the models.

The analysis was performed by N. K. S. and A. S. N. using the *survival* package and R version 4.2.2 (R Foundation for Statistical Computing) [13–15].

## RESULTS

Of 51982 eligible subjects, 965 (1.9%) were excluded because of missing age or gender. Of the remaining 51017 employees included, 3294 (6.5%) were censored during the study because of termination of employment. By the end of the study, 13134 (26%) had received the bivalent vaccine, which was the Pfizer vaccine in 11397 (87%) and the Moderna vaccine in the remaining 1700. Altogether, 4424 employees (8.7%) acquired COVID-19 during the 26 weeks of the study.

### Baseline characteristics

Table 1 shows the characteristics of subjects included in the study. Notably, this was a relatively young population, with a mean age of 42 years. Among these, 20686 (41%) had previously had a documented episode of COVID-19 and 13717 (27%) had previously had an Omicron variant infection. 45064 subjects (88%) had previously received at least one dose of vaccine, 42550 (83%) had received at least two doses, and 46761 (92%) had been previously exposed to SARS-CoV-2 by infection or vaccination.

**Table 1.**
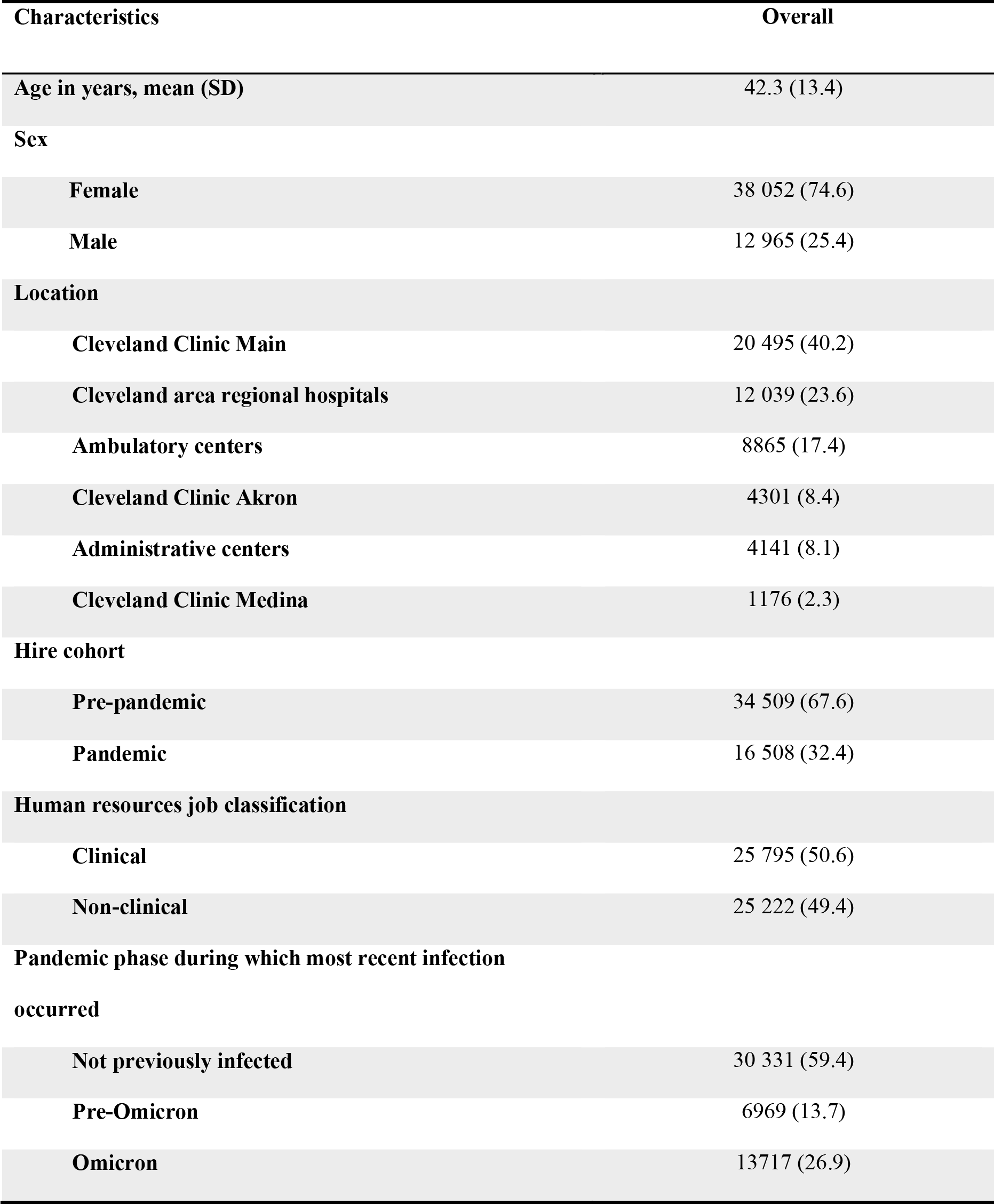
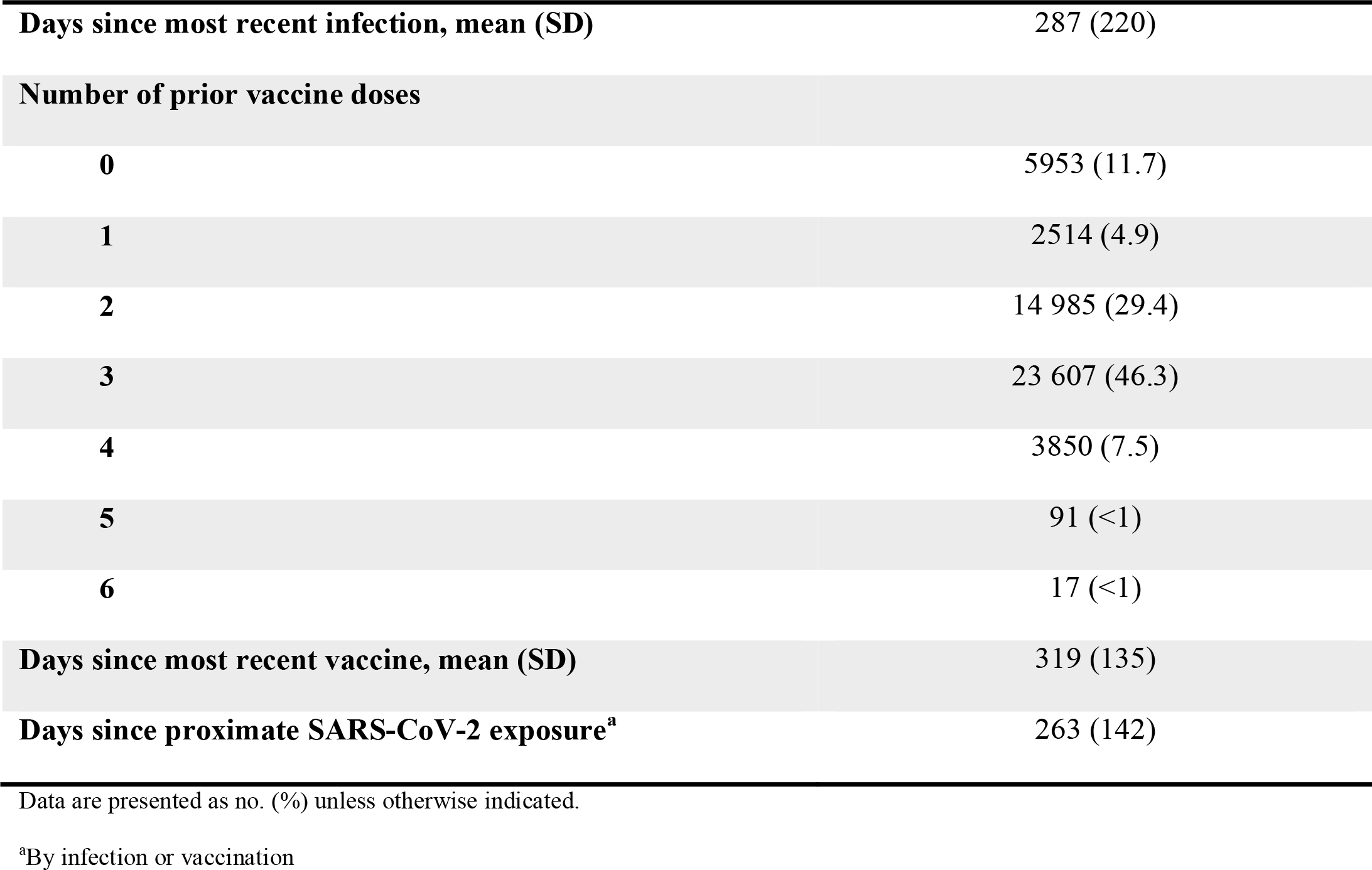
Baseline characteristics of 51017 employees of Cleveland Clinic in Ohio.

| Characteristics | Overall |
| --- | --- |
| Age in years, mean (SD) | 42.3 (13.4) |
| Sex |  |
| Female | 38 052 (74.6) |
| Male | 12 965 (25.4) |
| Location |  |
| Cleveland Clinic Main | 20 495 (40.2) |
| Cleveland area regional hospitals | 12 039 (23.6) |
| Ambulatory centers | 8865 (17.4) |
| Cleveland Clinic Akron | 4301 (8.4) |
| Administrative centers | 4141 (8.1) |
| Cleveland Clinic Medina | 1176 (2.3) |
| Hire cohort |  |
| Pre-pandemic | 34 509 (67.6) |
| Pandemic | 16 508 (32.4) |
| Human resources job classification |  |
| Clinical | 25 795 (50.6) |
| Non-clinical | 25 222 (49.4) |
| Pandemic phase during which most recent infection occurred |  |
| Not previously infected | 30 331 (59.4) |
| Pre-Omicron | 6969 (13.7) |
| Omicron | 13717 (26.9) |
| **Days since most recent infection, mean (SD)** | 287 (220) |
| **Number of prior vaccine doses** |  |
| **0** | 5953 (11.7) |
| **1** | 2514 (4.9) |
| **2** | 14 985 (29.4) |
| **3** | 23 607 (46.3) |
| **4** | 3850 (7.5) |
| **5** | 91 (<1) |
| **6** | 17 (<1) |
| **Days since most recent vaccine, mean (SD)** | 319 (135) |
| **Days since proximate SARS-CoV-2 exposure<sup>a</sup>** | 263 (142) |

Data are presented as no. (%) unless otherwise indicated.
<sup>a</sup>By infection or vaccination

### Risk of COVID-19 based on prior infection and vaccination history

The risk of COVID-19 varied by the phase of the epidemic in which the subject’s last prior COVID-19 episode occurred. In decreasing order of risk were those never previously infected, those last infected during the pre-Omicron phase, and those last infected during the Omicron phase (Figure 1).

**Figure 1.**
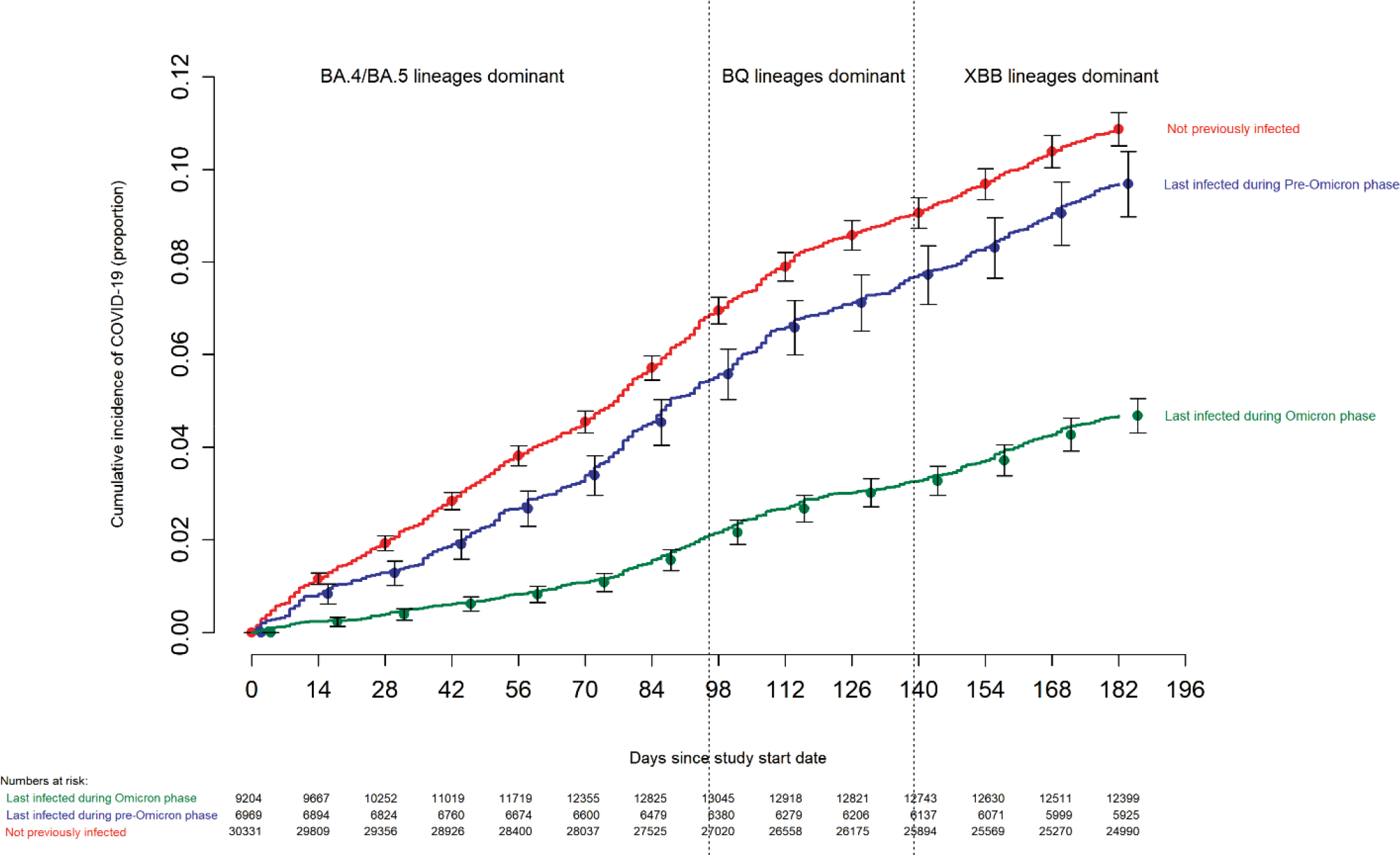
Cumulative incidence of COVID-19 for subjects stratified by the pandemic phase during which the subject’s last prior COVID-19 episode occurred. Day zero was 12 September 2022, the day the bivalent vaccine began to be offered to employees. Point estimates and 95% confidence intervals are jittered along the x-axis to improve visibility.

The risk of COVID-19 also varied by the number of COVID-19 vaccine doses previously received. The higher the number of vaccines previously received, the higher the risk of contracting COVID-19 (Figure 2).

**Figure 2.**
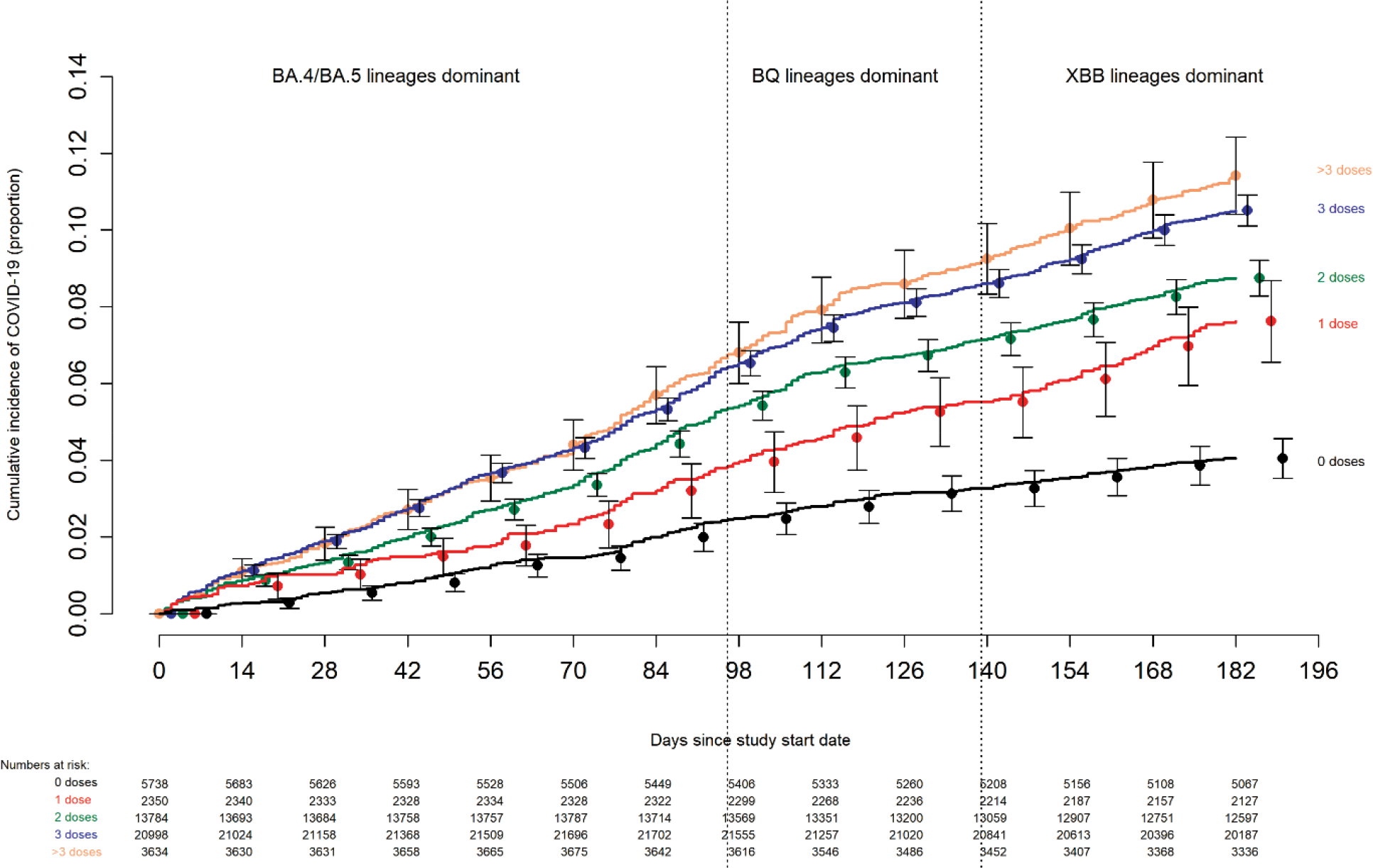
Cumulative incidence of COVID-19 for subjects stratified by the number of COVID-19 vaccine doses previously received. Day zero was 12 September 2022, the day the bivalent vaccine began to be offered to employees. Point estimates and 95% confidence intervals are jittered along the x-axis to improve visibility.

### Bivalent vaccine effectiveness

The cumulative incidence of COVID-19 was similar for the bivalent vaccinated and non-bivalent vaccinated states in an unadjusted analysis (Figure 3).

**Figure 3.**
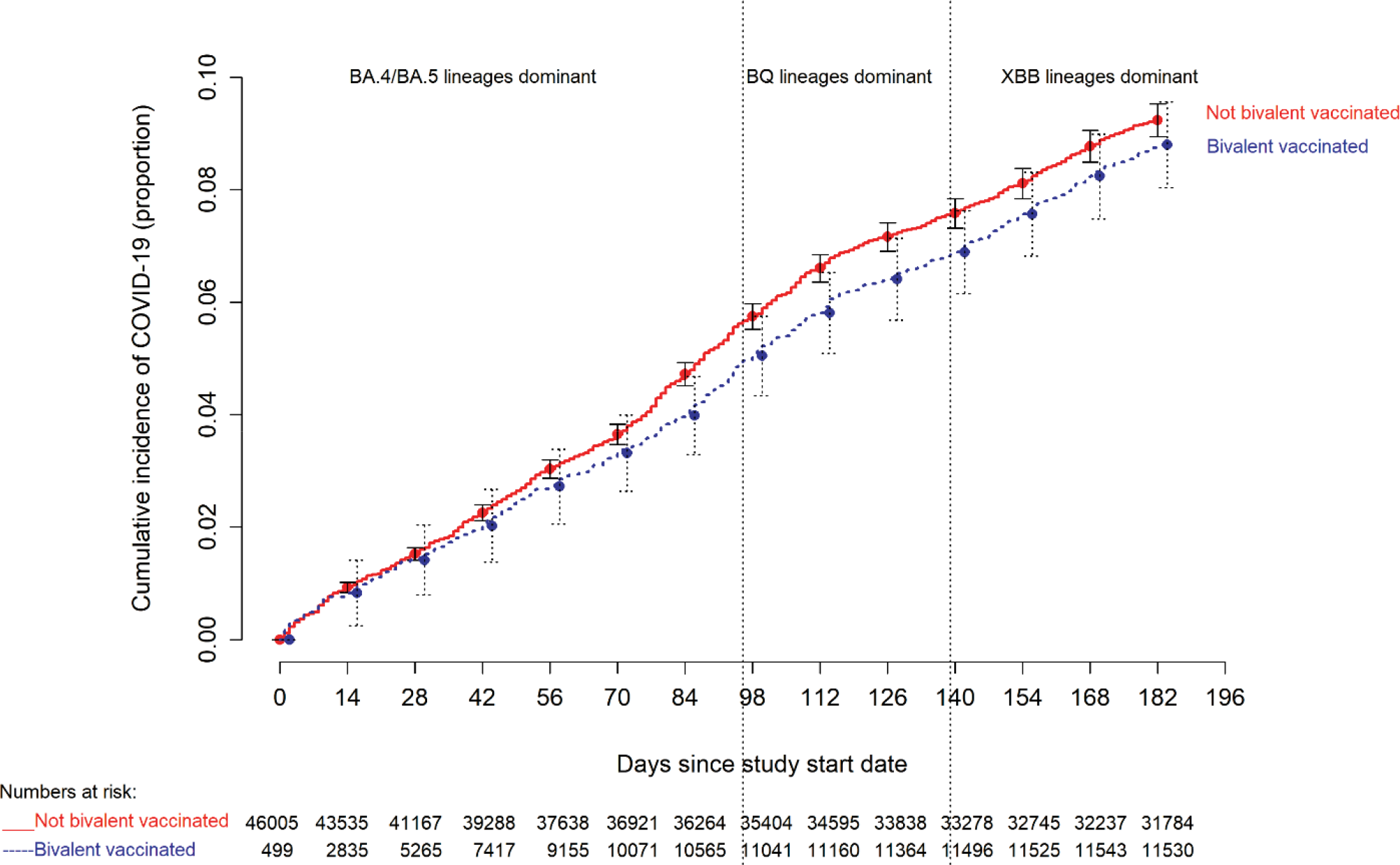
Simon-Makuch plot comparing the cumulative incidence of COVID-19 for the bivalent vaccinated and non-bivalent vaccinated states. Day zero was 12 September 2022, the day the bivalent vaccine began to be offered to employees. Point estimates and 95% confidence intervals are jittered along the x-axis to improve visibility.

In a multivariable Cox proportional hazards regression model, adjusted for age, gender, hire cohort, job category, number of COVID-19 vaccine doses prior to study start, and epidemic phase when the last prior COVID-19 episode occurred, bivalent vaccination provided some protection against COVID-19 while the BA.4/5 lineages were the dominant circulating strains (HR, .71; 95% C.I., .63-.79; P-value, <.001), and less protection while the BQ lineages were dominant (HR, .80; 95% C.I., .69-.94; P-value, .005). A protective effect of bivalent vaccination could not be demonstrated while the XBB strains were dominant (HR, 0.96; 95% C.I., .82-.1.12; P-value, .59). Point estimates and 95% confidence intervals for hazard ratios for the variables included in the unadjusted and adjusted Cox proportional hazards regression models are shown in Table 2. The calculated overall bivalent vaccine effectiveness from the model was 29% (95% C.I., 21%-37%) during the BA.4/5 dominant phase, 20% (95% C.I., 6%-31%) during the BQ dominant phase, and 4% (95% C.I., -12%-18%) during the XBB dominant phase.

**Table 2.**
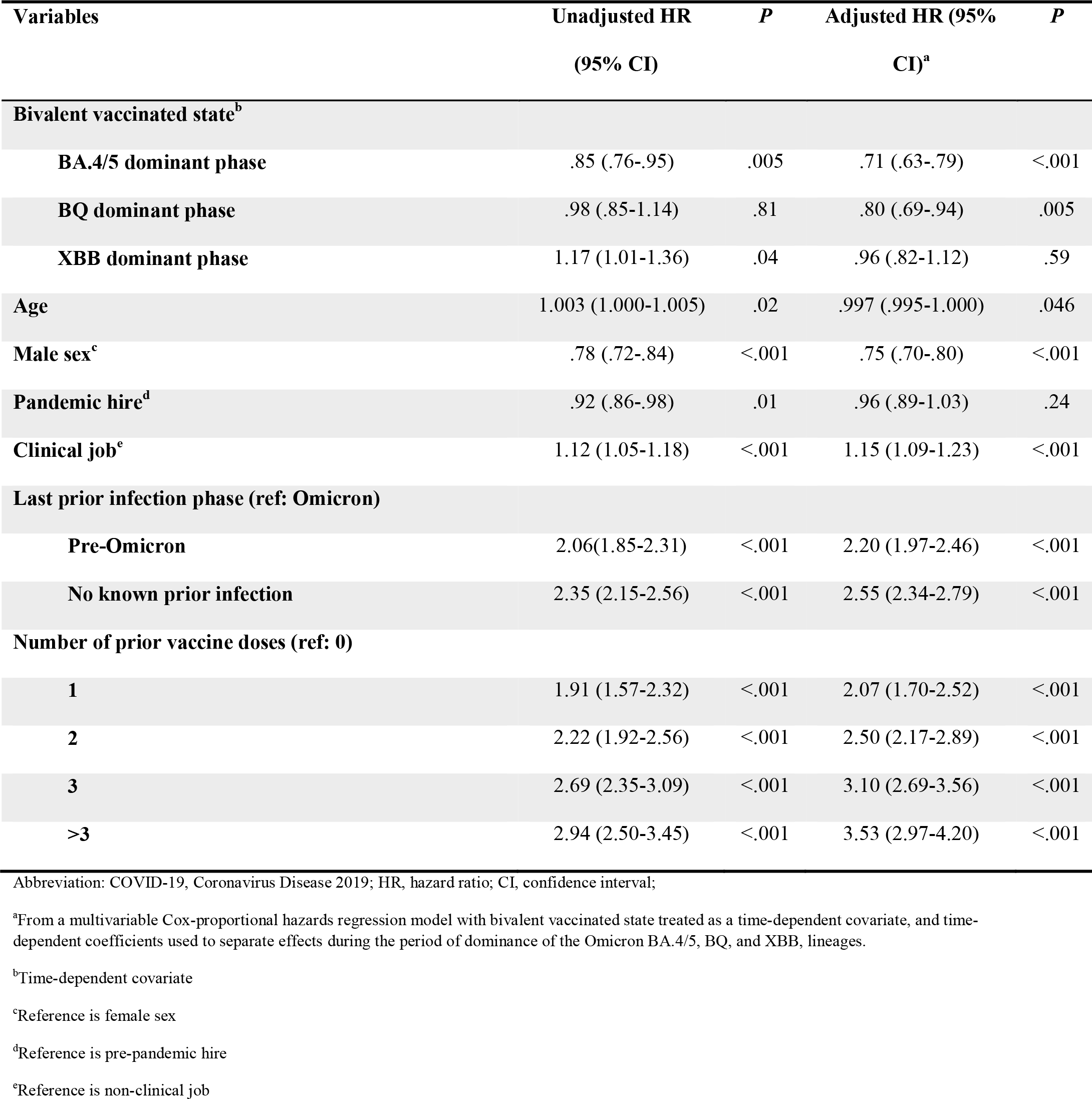
Unadjusted and Adjusted Associations with Time to COVID-19.

The multivariable analysis also found that, the more recent the last prior COVID-19 episode was the lower the risk of COVID-19, and that the greater the number of vaccine doses previously received the higher the risk of COVID-19.

### Bivalent vaccine effectiveness among those with prior SARS-CoV-2 infection or vaccination

Among persons with prior exposure to SARS-CoV-2 by infection or vaccination, hazard ratios for bivalent vaccination for individuals, after adjusting for time since proximate SARS-CoV-2 exposure, are shown in table 3. Bivalent vaccination protected against COVID-19 during the BA.4/5 dominant phase (HR, .78; 95% C.I., .70-.88; P-value, <.001), but a significant protective effect could not be demonstrated during the BQ dominant phase (HR, .91; 95% C.I., .78-.1.07; P-value, .25) or the XBB dominant phase (HR, 1.05; 95% C.I., .85-.1.29; P-value, .66).

**Table 3.**
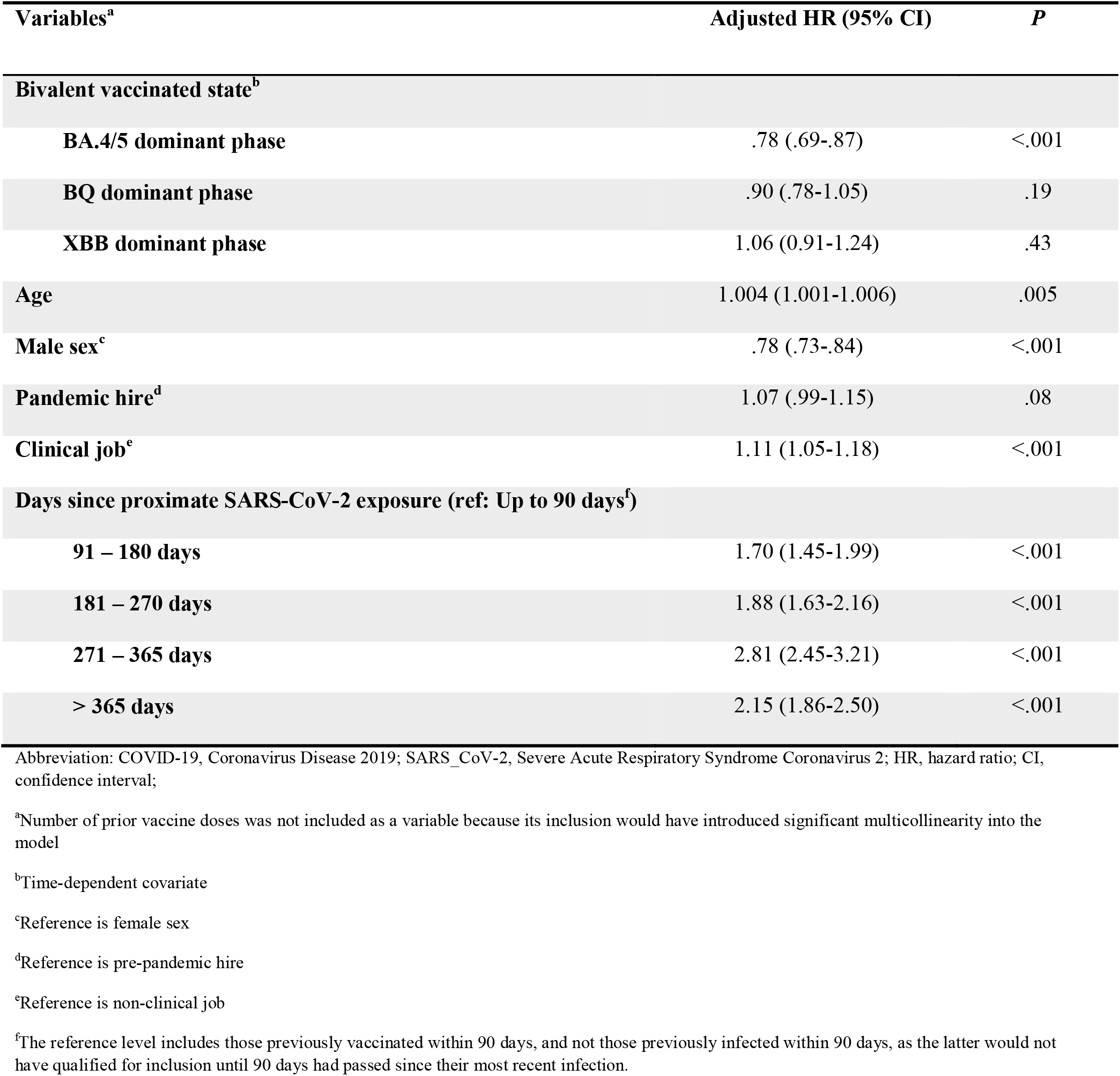
Adjusted associations with time to COVID-19, adjusted for time since proximate SARS-CoV-2 exposure by prior infection or prior vaccination, among those with prior SARS-CoV-2.

## DISCUSSION

This study found that the current bivalent vaccines were about 29% effective overall in protecting against infection with SARS-CoV-2 when the Omicron BA.4/5 lineages were the predominant circulating strains, and effectiveness was lower when the circulating strains were no longer represented in the vaccine. A protective effect could not be demonstrated when the XBB lineages were dominant. The magnitude of protection afforded by bivalent vaccination while the BA.4/5 lineages were dominant was similar to that estimated in another study using data from the Increasing Community Access to Testing (ICATT) national SARS-CoV-2 testing program [16].

The strengths of our study include its large sample size, and its conduct in a healthcare system where a very early recognition of the critical importance of maintaining an effective workforce during the pandemic led to devotion of resources to have an accurate accounting of who had COVID-19, when COVID-19 was diagnosed, who received a COVID-19 vaccine, and when. The study methodology, treating bivalent vaccination as a time-dependent covariate, allowed for determining vaccine effectiveness in real time.

The study has several limitations. Individuals with unrecognized prior infection would have been misclassified as previously uninfected. Since prior infection protects against subsequent infection, such misclassification would have resulted in underestimating the protective effect of the vaccine. However, there is little reason to suppose that prior infections would have been missing in the bivalent vaccinated and non-vaccinated states at disproportionate rates. There might be concern that those who chose to receive the bivalent vaccine might have been more worried about infection and might have been more likely to have got tested when they had symptoms, thereby disproportionately detecting more incident infections among those who received the bivalent vaccine. We did not find an association between the number of COVID-19 tests done and number of prior vaccine doses, suggesting that this was not a confounding factor. Those who chose to get the bivalent vaccine would have been those who were more likely to have lower risk-taking behavior with respect to COVID-19. This would have the effect of finding a higher risk of COVID-19 in the non-vaccinated state (as those who chose not to get the bivalent vaccine, expectedly with higher risk-taking behavior, remained in the non-vaccinated state throughout the duration of the study), thereby potentially overestimating vaccine effectiveness. The widespread availability of home testing kits might have reduced detection of incident infections. This potential effect should be somewhat mitigated in our healthcare cohort because one needs a NAAT to get paid time off, providing a strong incentive to get a NAAT if one tests positive at home. Even if one assumes that some individuals chose not to follow up on a positive home test result with a NAAT, it is very unlikely that individuals would have chosen to pursue NAAT after receiving the bivalent vaccine more so than before receiving the vaccine, at rates disproportionate enough to affect the study’s findings. We were unable to distinguish between symptomatic and asymptomatic infections, and had to limit our analyses to all detected infections. Variables that were not considered might have influenced the findings substantially. There were too few severe illnesses for the study to be able to determine if the vaccine decreased severity of illness. Lastly, our study was done in a healthcare population, and included no children and few elderly subjects, and the majority of study subjects would not have been immunocompromised.

A possible explanation for a weaker than expected vaccine effectiveness is that a substantial proportion of the population may have had prior asymptomatic Omicron variant infection. About a third of SARS-CoV-2 infections have been estimated to be asymptomatic in studies that have been done in different places at different times [17–19]. If so, protection from the bivalent vaccine may have been masked because those with prior Omicron variant infection may have already been somewhat protected against COVID-19 by virtue of natural immunity. A seroprevalence study conducted by the CDC found that by February 2022, 64% of the 18-64 age-group population and 75% of children and adolescents had serologic evidence of prior SARS-CoV-2 infection [20], with almost half of the positive serology attributed to infections that occurred between December 2021 and February 2022, which would have predominantly been Omicron BA.1/BA.2 lineage infections. With such a large proportion of the population expected to have already been previously exposed to the Omicron variant of SARS-CoV-2, there could be some concern that a substantial proportion of individuals may be unlikely to derive any meaningful benefit from a bivalent vaccine.

The association of increased risk of COVID-19 with higher numbers of prior vaccine doses was unexpected. A simplistic explanation might be that those who received more doses were more likely to be individuals at higher risk of COVID-19. A small proportion of individuals may have fit this description. However, the majority of subjects in this study were generally young individuals and all were eligible to have received at least 3 doses of vaccine by the study start date, and which they had every opportunity to do. Therefore, those who received fewer than 3 doses (46% of individuals in the study) were not those ineligible to receive the vaccine, but those who chose not to follow the CDC’s recommendations on remaining updated with COVID-19 vaccination, and one could reasonably expect these individuals to have been more likely to have exhibited higher risk-taking behavior. Despite this, their risk of acquiring COVID-19 was lower than those who received a larger number of prior vaccine doses. This is not the only study to find a possible association with more prior vaccine doses and higher risk of COVID-19. During an Omicron wave in Iceland, individuals who had previously received 2 or more doses were found to have a higher odds of reinfection than those who had received fewer than 2 doses of vaccine, in an unadjusted analysis [21]. A large study found, in an adjusted analysis, that those who had an Omicron variant infection after previously receiving three doses of vaccine had a higher risk of reinfection than those who had an Omicron variant infection after previously receiving two doses of vaccine [22]. Another study found, in multivariable analysis, that receipt of two or three doses of a mRNA vaccine following prior COVID-19 was associated with a higher risk of reinfection than receipt of a single dose [7]. Immune imprinting from prior exposure to different antigens in a prior vaccine [22,23], and class switch towards non-inflammatory spike-specific IgG4 antibodies after repeated SARS-CoV-2 mRNA vaccination [24], have been suggested as possible mechanisms by why prior vaccine may provide less protection than expected. We still have a lot to learn about protection from COVID-19 vaccination, and in addition to a vaccine’s effectiveness, it is important to examine whether multiple vaccine doses given over time may not be having the beneficial effect that is generally assumed.

In conclusion, this study found an overall modest protective effect of the bivalent vaccine against COVID-19 while the circulating strains were represented in the vaccine and lower protection when the circulating strains were no longer represented. A significant protective effect was not found when the XBB lineages were dominant. The unexpected finding of increasing risk with increasing number of prior COVID-19 vaccine doses needs further study.

## Data Availability

All data produced in the present study are available upon reasonable request to the authors

## Notes

### Author contributions

N. K. S.: Conceptualization, methodology, validation, investigation, data curation, software, formal analysis, visualization, writing-original draft preparation, writing-reviewing and editing, supervision, project administration. P. C. B.: Resources, investigation, validation, writing-reviewing and editing. A. S. N.: Methodology, formal analysis, visualization, validation, writing-reviewing and editing. J. F. S.: Resources, writing-reviewing and editing. A. H.: Resources, writing-reviewing and editing. S. M. G.: Project administration, resources, writing-reviewing and editing.

### Potential conflicts of interest

The authors: No reported conflicts of interest. All authors have submitted the ICMJE Form for Disclosure of Potential Conflicts of Interest. Conflicts that the editors consider relevant to the content of the manuscript have been disclosed.

### Funding

None.

## Notes

### Competing Interest Statement

The authors have declared no competing interest.

### Funding Statement

This study did not receive any funding

### Author Declarations

The Institutional Review Board of Cleveland Clinic gave ethical approval for this work.

### Summary of Updates

The only difference from the immediate preceding version is that Figure legends for figures 1 and 2 were corrected. The data for this study were obtained from a living database. Vaccine information for vaccines received outside the Cleveland Clinic Health System populate the database when these data become available from other health systems or from the Ohio Department of Health. Additionally errors are corrected when identified. Because of these circumstances, data pulled on subsequent dates are more complete than those pulled earlier. Occasionally data on individual patients may differ from those pulled earlier because errors previously present have been corrected. The findings of this version of the study are based on data extracted on 2/21/2023.

